# *Clostridioides difficile* Detection in a Human CRC Cohort

**DOI:** 10.64898/2026.02.20.26346702

**Authors:** SM Anderson, Z Cing, JL Drewes, JR White, T Southward, H Beauregard, JT Ferri, JW Wanyiri, A Roslani, J Vadivelu, SN Tang, J Queen, CL Sears

**Affiliations:** Johns Hopkins University School of Medicine, Baltimore, MD, USA; University of Maryland Baltimore County, Baltimore, MD, USA; Resphera Biosciences, Baltimore, MD, USA; Universiti Malaya, Kuala Lumpur, Malaysia

## Abstract

**Background:** The role of the gut microbiome and specific enteric bacteria in influencing the development of colorectal cancer (CRC) remains incompletely understood. Recently, it was shown that human CRC-derived strains of *Clostridioides difficile* were capable of inducing colonic tumorigenesis in a susceptible mouse model. We hypothesized that *C. difficile* contributes to the pathogenesis of human CRC and would be enriched in CRC tumors compared to paired normal tissues from the same individual.

**Methods:** We analyzed matched tumor/normal tissue samples from a cohort of 108 individuals presenting to a tertiary care hospital in Kuala Lumpur, Malaysia for CRC resection between 2013-2014. We assessed the prevalence of *C. difficile* detection using 16S rRNA amplicon sequencing with high-resolution taxonomic assignment as well as culture and PCR.

**Results:** We found that detection of *C. difficile* was prevalent (38% of individuals), but of low abundance (tumor median relative abundance 0.01%, paired normal 0.006% [p=0.4]). Detection of *C. difficile* was more prevalent in individuals with biofilm-positive tumor tissues than biofilm-negative (i.e., 81% of *C. difficile*-positive individuals were biofilm-positive vs. 63% of *C. difficile*-negative individuals [p=0.04]). Additionally, in exploratory analyses, we describe patterns of taxonomic and inferred functional pathway differences between *C. difficile-*positive and *C. difficile*-negative groups.

**Conclusion:** These findings suggest that *C. difficile* is frequently present in low abundance in the tumor microbiome with a potentially significant impact on community composition and function.

## BACKGROUND

Sporadic colorectal cancer (CRC) is a significant global public health concern, notable for a rising incidence of early-onset cases, now the leading cause of cancer deaths in young individuals (EO-CRC, i.e., age <50) (1, 2). The gut microbiome has been shown to contribute to CRC pathogenesis, but these interactions are complex and incompletely understood (3). Next generation sequencing methods (e.g., 16S ribosomal RNA [rRNA] amplicon sequencing) have been extensively deployed to identify and characterize microbes present in stool or colonic tissue, advancing our understanding of the associations between the gut microbiome and the natural history of colorectal neoplasia. Prior research has identified both broad microbiota signatures (4) and individual gut bacteria (5-7) associated with CRC pathogenesis. Importantly, many of these studies report enriched or depleted relative abundance of bacterial taxa or operational taxonomic units as primary outcome measures in an attempt to quantify their importance to CRC development.

A new member was recently added to the list of potential individual bacterial contributors to CRC when it was shown that human CRC-derived strains of toxigenic *Clostridioides difficile* could induce colonic tumorigenesis in a susceptible mouse model (Min or multiple intestinal neoplasia mice heterozygous for *Apc*) (8). Notable nuances to this finding included the necessity of persistent toxin B (TcdB) production (but not TcdA or binary toxin) *in vivo* to affect tumorigenesis and the ability of *C. difficile* to promote tumorigenesis despite a low relative abundance (<2%) compared to other microbes. These unexpected observations led us to hypothesize that *C. difficile* functions as a keystone species (i.e., one that exerts a disproportionately high ecological influence relative to its abundance) and thus may be an unrecognized microbe contributing to changes in CRC epidemiology.

To address this hypothesis, we analyzed a cohort of individuals from Malaysia (N=108) who presented for surgical resection of CRC. Tumors and paired normal tissues from the surgical resection margin were analyzed with high-resolution 16S rRNA amplicon sequencing and molecular- and culture-based methods to define the prevalence of *tcdB+ C. difficile* (hereafter, toxigenic *C. difficile*) in this cohort.

## METHODS

### Study Participants

This study was approved by the Johns Hopkins Institutional Review Board and the University of Malaya Medical Centre (UMMC, Kuala Lumpur, Malaysia) Medical Ethics Committee. Individuals ≥18 years old with a diagnosis of CRC were eligible for enrollment. Written informed consents (provided in Malay and English) were obtained from all participants. All participants were enrolled in accordance with the Health Insurance Portability and Accountability Act. Samples were collected prospectively during the years 2013-2014. Individuals who had received pre-operative radiation, chemotherapy or had a personal prior history of CRC were excluded. All individuals in the study underwent standard mechanical bowel preparation. Standard pre-operative intravenous antibiotics were administered in all surgical cases but oral antibiotics were not standard of care before surgery at UMMC at the time of the study. We collected data on the following demographic variables: age, sex, ethnicity, body mass index (BMI), smoking history, and alcohol consumption, and the following tumor-intrinsic variables: tumor location, tumor pathology, tumor grade, and tumor stage. Individuals enrolled under age 50 were defined as having early-onset CRC (EO-CRC), whereas individuals aged 50 and over were defined as having late-onset CRC (LO-CRC). Study data were collected and managed using REDCap electronic data capture tools hosted at Johns Hopkins University.

### Sample Collection

On the day of surgery, excess colon tumor and paired normal tissues (from the most distal edge of the surgical resection) not needed for pathologic clinical analyses, were collected for laboratory analyses. Surgical tissues were fixed in methacarn or flash frozen (-80°C) for later analysis as previously published (9).

### Illumina 16S rRNA Amplicon Gene Library Generation, Sequencing and Analysis

16S rRNA amplicon sequencing of the 108 tumor-normal paired tissues was performed, over time, at different facilities with different methodologies yielding 3 sequencing sub-cohorts, MAL1, MAL2 and MAL3 as previously described (9) (**Table S1**). For each cohort, the V3-V4 hypervariable region of the 16S rRNA gene was amplified and sequenced.

Paired-end 16S rRNA amplicon sequences were trimmed for quality and length as previously described (9). High-quality 16S rRNA amplicon sequences were assigned to a high-resolution taxonomic lineage using Resphera Insight, which uses a hybrid global-local alignment strategy to a manually curated 16S rRNA database with 11,000 unique species (4, 10, 11). This approach attempts to achieve species-level resolution when possible; however, when a confident single species assignment is not feasible, the method minimizes false positives by providing “ambiguous assignments” i.e., a list of candidate species reflecting the ambiguity. Taxonomic assignments are presented as unambiguous assignments by Resphera Insight unless otherwise indicated. Normalization of observations was performed by rarefaction (rarefaction levels as stated in Results) followed by alpha and beta diversity characterization. Taxonomic abundances were converted from counts to percentages of passable reads analyzed per sample. Functional inference of microbial gene content was evaluated using PICRUSt2 (12).

16S rRNA amplicon sequencing was considered positive for *C. difficile* if at least one confident *C. difficile* read was detected. Use of this low level of read number to define *C. difficile* positivity is based on two studies identifying that a single read of *C. difficile* in a human sample has been verified as positive when inoculated into mice where *C. difficile* ‘blooms’ in the murine feces (8, 13).

### Limit of Detection Analysis

The expected limit of detection of *C. difficile* for each sequencing sample was calculated using the q-beta function in R to generate a limit of detection (LOD) curve by modeling detection probability across varying relative abundances, assuming a beta distribution reflective of observed sequencing depth and prevalence. This approach enabled visualization of the detection threshold at a defined confidence level (p = 0.05) within each tissue sample analyzed.

### Selective Culture and Identification of *C. difficile* from Samples

3mm punch biopsies were collected from frozen tumor and normal tissue, mechanically homogenized, and resuspended for targeted culture of *C. difficile* as further described in supplemental methods. Species identification of *C. difficile* was confirmed by Sanger sequencing of the full-length 16S rRNA gene (Azenta Life Sciences) for at least one colony from each individual who yielded a positive culture result. Methods for UMMC culturing from stool samples are available in the supplemental methods section.

### PCR for Toxin B Assay

*C. difficile* strains isolated by culture were analyzed for the presence of toxin genes (including *tcdA, tcdB, cdtA, cdtB*) utilizing a pan-toxin PCR as previously described (14). Details are available in supplemental methods.

### Quantification and statistical analysis

To compare categorical data between defined groups (e.g., *C. difficile* status by biofilm presence), data were analyzed using the Chi-Squared Test. Continuous demographic variables were compared using an unpaired t-test. For comparisons of relative taxonomic abundances between individuals displaying *C. difficile* and those in which *C. difficile* was not detected, data were analyzed by the Mann-Whitney test. Differences in PICRUSt2 pathway content was also analyzed by Mann Whitney test, as indicated in the figure legends. For all analyses, differences with a p-value of <0.05 were considered significant.

Methods for FISH analysis of biofilms and the data availability statement are discussed in supplemental methods.

## RESULTS

### Detection of *C. difficile* in CRC and Paired Normal Samples by High-Resolution 16S rRNA Amplicon Sequencing and High-Resolution Taxonomic Assignment

108 individuals had matching tumor and normal colonic tissue collected and preserved for analysis in this study. From both tumor and paired normal samples from these individuals, 16S rRNA amplicon sequence data sets were analyzed using Resphera Insight, a high-resolution methodology for species-level characterization with established sensitivity and specificity for the detection of *C. difficile* (11). *C. difficile* at any abundance was detected in 28/108 (26%) of tumor biopsies and 25/108 (24%) of paired normal biopsies (**Fig 1a**). Combined, forty-one individuals (38%) had *C. difficile* detected in either tumor or normal tissue. Twelve individuals (11%) had *C. difficile* detected in both tumor and normal samples. The relative abundance of *C. difficile* in all detected samples was low, with a median of 0.01% (IQR 0.006-0.02%) in the tumor samples and 0.006% (IQR 0.005-0.02%) in the paired normal margin samples (p=0.4, **Fig 1b**).

**Figure 1.**
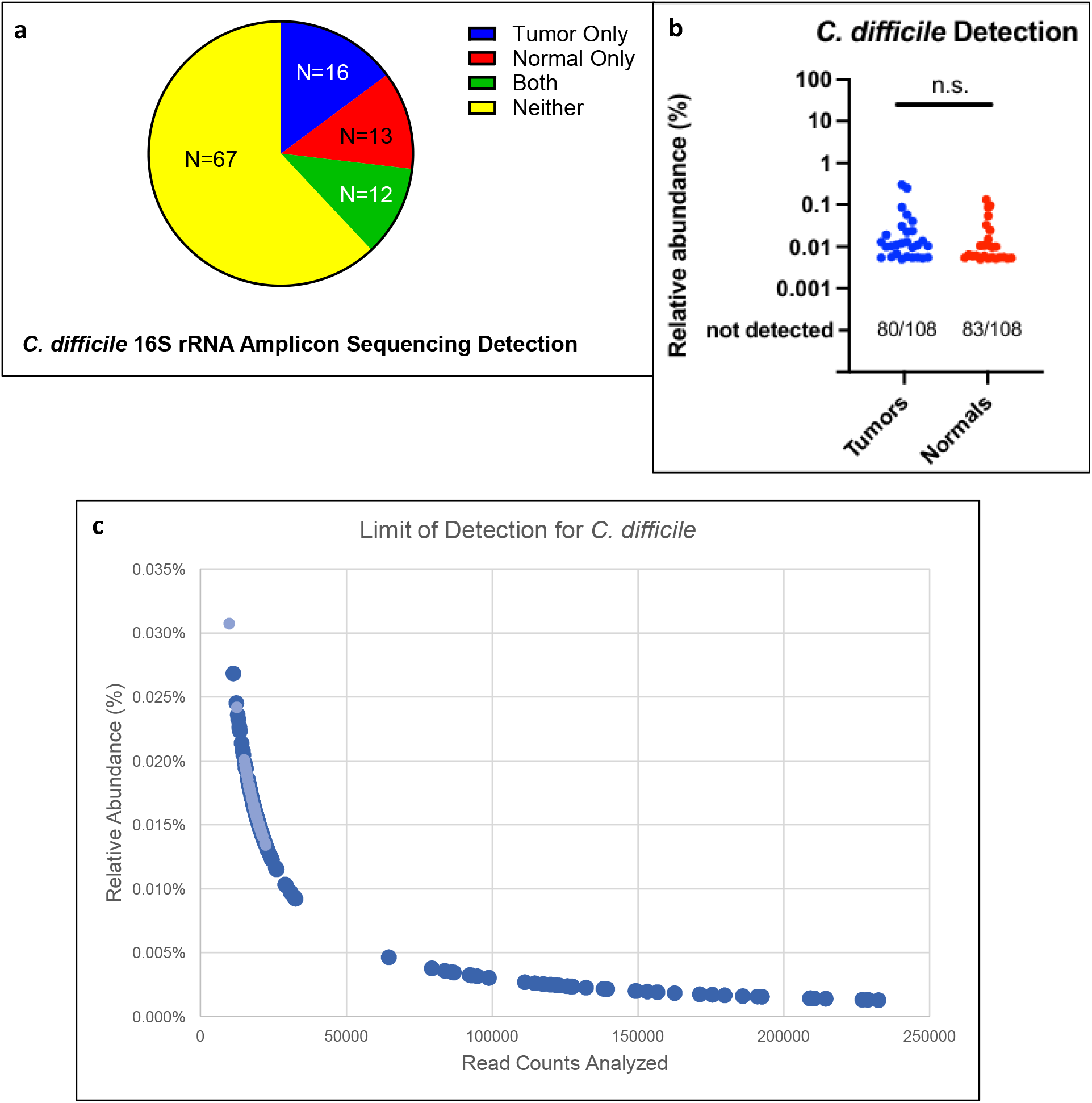
*C. difficile* Detection by 16S rRNA Amplicon Sequencing. **a)***C. difficile* detection (>0 reads) by 16S rRNA amplicon sequencing and high-resolution taxonomic assignment stratified by the tissue type in which it was detected for each individual. N lists the number of unique individuals with CD detection by tissue type. **b)** Relative abundance of *C. difficile* as detected by 16S rRNA amplicon sequencing in each tissue; median values are not significantly different by paired Wilcoxon rank-sum test. **c)** Limit of detection analysis as calculated from our data to estimate the read depth needed to detect at least one amplicon read assigned to *C. difficile* at a given relative abundance of the organism, with 95% confidence. Light blue dots indicate samples with actual detection of *C. difficile* by 16S rRNA amplicon sequencing in our cohort. This figure is partially quantified in Supplemental Table 2.

### Analysis of Read Depth Effects

The initial 16S rRNA amplicon sequencing of the tissue samples from this cohort was performed in three sequencing runs (MAL1, MAL2, MAL3) on different sequencing platforms at differing times (see Methods and **Table S1**); this resulted in a heterogeneous quantity of passable sequencing reads available for analysis, with a median of 19,501 reads per sample and a range from 9,738 to 232,430 reads. When applying a universal cutoff of 9500 random reads for analysis to allow for equitable inclusion of each sample, the sensitivity for *C. difficile* detection fell by 42% (from 41 positive individuals to 24, **Table 1**). Using our data and the q-beta function of R, we calculated the expected limit of detection for *C. difficile* by modeling detection probability across varying relative abundances, demonstrating that read depth has a marked effect on the detection of low-abundance *C. difficile* in bacteria-dense colon tissue samples (**Fig 1c; Table 1**).

**Table 1.**
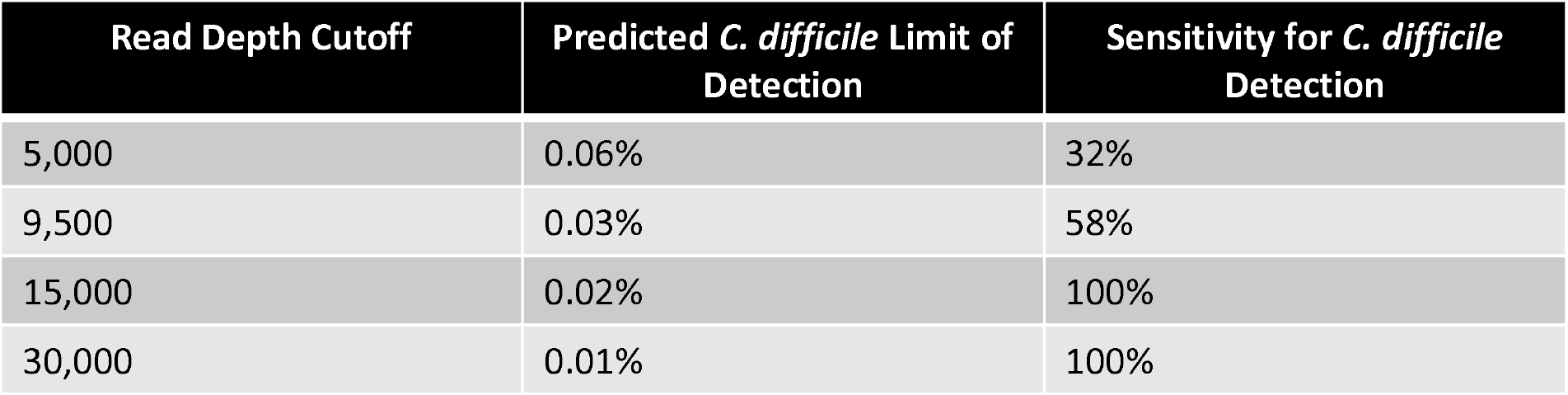
Limit of Detection and *C. difficile* Detection Sensitivity. Using the Limit of Detection Analysis calculation described in Methods, we list the expected relative abundance of *C. difficile* at benchmark read depth cutoffs above which there is a 95% likelihood of detection of at least one read assigned as *C. difficile* by 16S rRNA amplicon sequencing. The third column provides the estimated sensitivity for any *C. difficile* detection in all the individuals in this cohort at these benchmark read depth cutoffs using our unrestricted analysis (median of 19,501 reads per sample) as the gold standard.

### Isolation of *C. difficile* by Selective Culturing and Toxin B Analysis of Tumor and Normal Tissue Biopsies

To validate the sequencing findings, tumor and normal tissue samples from individuals identified as *C. difficile-*positive by 16S rRNA amplicon sequencing were cultured for isolation of *C. difficile* strains. Forty-one individuals had at least one 16S rRNA amplicon sequence read assigned to *C. difficile* in either tumor or normal samples (**Fig 1a**); forty of these individuals had both tumor and normal tissue available for culture. To complement these cultures, tumor tissues from 25 randomly selected individuals without any sequencing reads assigned to *C. difficile* were also cultured.

*C. difficile* was recovered by culture from either tumor or normal tissue in 9 of 40 individuals (23%) identified as *C. difficile-*positive by 16S rRNA amplicon sequencing and in 1 of 25 (4%) tumor tissue samples from individuals who had no 16S rRNA amplicon reads assigned to *C. difficile* in tumor or normal tissue (**Table 2a**). The culture success rate was greater for tissue-concordant samples (i.e., positive tumor culture for individuals with positive *C. difficile* 16S rRNA amplicon sequencing of tumor tissue): 21% *C. difficile* culture-positive from sequence-positive tissue vs. 5.7% culture-positive from sequence-negative tissue, p=0.02) (**Table 2b**).

**Table 2.** Cohort Microbiologic Features & Tissue Sequencing-Culture Concordance. **a)**Results of targeted culturing of tissue samples for *C. difficile* stratified by tissue 16S rRNA amplicon sequencing classification (top row labels) and tissue culture isolation of *C. difficile* (first column labels). Underlined fractions / percentages are considered “concordant” (i.e., a <u>tissue</u> with detection of *C. difficile* by 16S rRNA amplicon sequencing also yielding a positive culture); data used for further analysis in Table 3B. **b)** Concordance or discordance is shown for *C. difficile* 16S rRNA amplicon-sequencing compared to *C. difficile*-directed culture of each individual <u>tissue</u>. For more details on the culture-positive individuals, see Supplemental Table 2. P-value determined by Chi-Squared Test. “Seq+” = 16S rRNA amplicon sequencing detection of at least one read assigned to *C. difficile* in the specified tissue sample.

|  | Only Tumor Seq+<br>CD (n=15 persons) | Only Normal Seq+<br>CD (n=13 persons) | Both Seq+ CD<br>(n=12 persons) | Both Seq- CD<br>(n=25 persons) |
| --- | --- | --- | --- | --- |
| **Tumor Culture+** | <u>4/15 (27%)</u> | 1/13 (8%) | <u>3/12 (25%)</u> | 1/25 (4%) |
| **Normal Culture+** | 1/15 (7%) | <u>2/13 (15%)</u> | <u>2/12 (17%)</u> | -- |

Tissue Sequencing-Culture Concordance Analysis
|  | Culture Positivity Rate for CD per Tissue Sample | P-Value |
| --- | --- | --- |
| Tissue Culture+/Seq+ (Concordant) | <u>11/52 (21%)</u> | 0.02* |
| Tissue Culture+/Seq- (Discordant) | 3/53 (5.7%) |  |

Underline: sequencing positive tissue, so a positive culture indicates concordance

Of the 10 individuals culture-positive for *C. difficile* from one or more tissues, the *tcdB* gene was detected via pan-toxin PCR in 7 of these individuals (**Table S2**).

### Demographic and Clinical Characteristics of *C. difficile* Positive Individuals and Association of *C. difficile* with CRC Biofilms

We assessed relevant demographic and clinical information for the 108 individuals (**Table 3**) to compare two groups: 1) those with tumor and/or paired normal samples positive for *C. difficile* by 16S rRNA amplicon sequencing (using all available passable reads) <u>or</u> with confirmed culture recovery of *C. difficile* (*C. difficile* positive subcohort, n=42 persons) and 2) those with tumor and paired normal samples negative for *C. difficile* (*C. difficile* negative subcohort, n=66 persons: 24 screened by both culture and sequencing, 42 screened by sequencing alone). Overall, we did not identify any demographic or tumor intrinsic factors significantly associated with detection of *C. difficile* in tissue biopsies. However, the *C. difficile* positive cohort had a significantly higher percentage of tumors classified as biofilm-positive by FISH (81% of *C. difficile* positive individuals with biofilms identified on their tumors vs. 63% of *C. difficile* negative individuals).

**Table 3.**
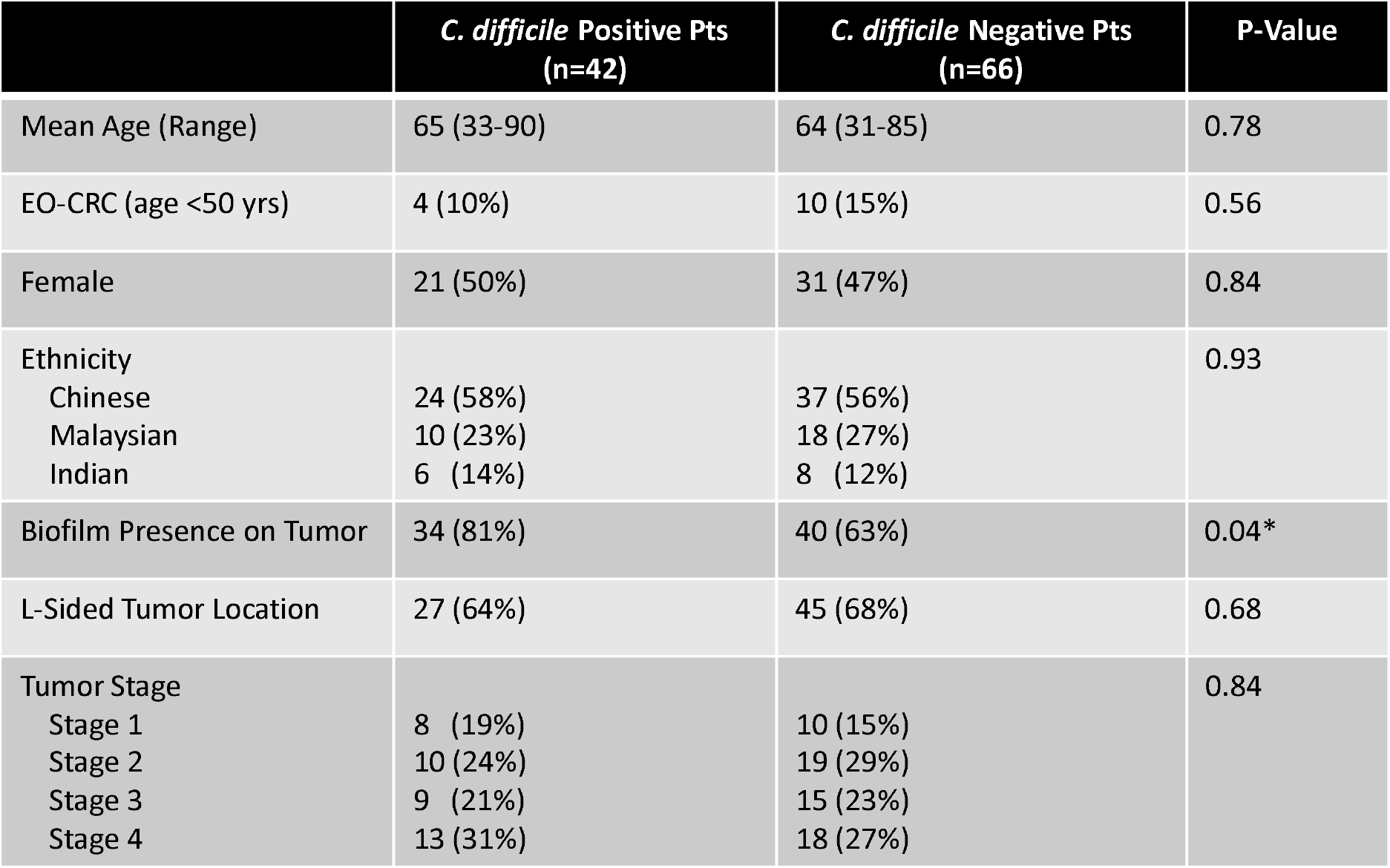
Cohort Demographic & Clinical Characteristics. Key clinical and demographic features compared between the *C. difficile* positive (defined as an individual having any tissue positive for *C. difficile* by 16S rRNA amplicon sequencing or culture) and *C. difficile* negative groups. P-values determined by unpaired t-test for continuous variables, Chi-Squared test for proportions.

### Microbiome Signatures of *C. difficile*-positive and -negative Individuals

Because in mice, low level *C. difficile* colonization leads to marked changes in the structure of biofilms as well as *C. difficile* and microbiota invasion into colonic crypts (8), we hypothesized that toxigenic *C. difficile* impacts human colon bacterial community structure and function even when present at very low relative abundance. Thus, we analyzed differences in *C. difficile*-positive or -negative community taxonomic structure as well as predicted functional pathway expression using the PICRUSt2 computational pipeline (12). Biofilms are associated with altered microbiome architecture (4, 9), thus, we removed biofilm status as a potential confounder by analyzing only biofilm-positive tumor samples to identify *C. difficile-*positive and *C. difficile*-negative associations.

We identified over 50 bacterial genera with significantly differential relative abundance between the *C. difficile*-positive and *C. difficile*-negative groups; **Fig 2** highlights some of the genera relevant to *C. difficile* biology and CRC pathogenesis, including several genera of Enterobacteriaceae (e.g., *Klebsiella, Enterobacter*) and Firmicutes (e.g., *Enterococcus, Lactobacillus*). Additionally, PICRUSt2 analysis suggested several bacterial functional pathways with significant differential abundance between the *C. difficile-*positive and *C. difficile*-negative subgroups (**Fig S1, Table S3**) including pathways consistent with putative carbohydrate degradation and the synthesis of secondary metabolites.

**Figure 2.**
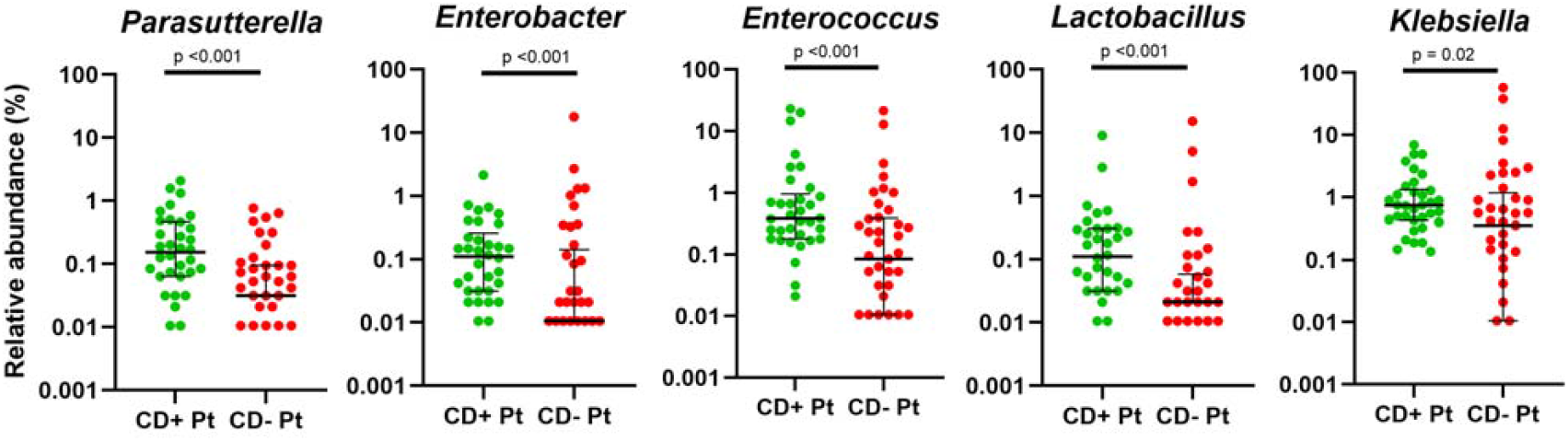
Community Signatures of *C. difficile*-Positive Individuals. Five bacterial genera of interest (relative abundances) were found to be significantly increased in *C. difficile* positive, biofilm positive (CD+ Pt) tumors versus *C. difficile* negative, biofilm positive (CD-Pt) tumors. P-values are determined by two-tailed non-parametric Wilcoxon signed-rank sum test

### *C. difficile* Culture-Positivity in non-CRC Malaysian Individuals

At UMMC where the surgical cohort was recruited, we obtained data from the clinical microbiology laboratory where *C. difficile* culture followed by TcdA and TcdB enzyme immunoassays (EIAs) on culture-positive samples was performed for physician-ordered *C. difficile* testing. Data were available from 2022-2023 (records from 2013-2014 were not available). During this time period, 119/2056 (5.8%) of all stools sent for *C. difficile* testing were culture-positive for *C. difficile*. 35 of these 119 isolates (29%) were positive for *C. difficile* toxin A and/or B byEIA (**Table S4**). Thus, overall, only 1.7% of stools sent for *C. difficile* testing at UMMC during this time period were positive for toxigenic *C. difficile*.

## DISCUSSION

In this cohort of individuals with CRC presenting to a Malaysian hospital for surgical tumor resection, we found an unexpectedly high prevalence of mucosal *C. difficile* (38% of individuals positive) in either tumor or paired normal tissue by 16S rRNA amplicon sequencing. This contrasts with the detection of *C. difficile* in only 5.8% of stool samples (1.7% for toxigenic *C. difficile*) sent for testing at this same hospital in the years 2022-2023. There is also a paucity of Malaysian literature identifying *C. difficile* as a prevalent enteric pathogen, either at the time of the study (2013-2014) or presently. However, other studies suggest similar rates of toxin B detection in the stool of hospitalized patients with diarrhea in Southeast Asia compared to Western countries (∼15-20% test positivity rate) (15). While our recent data (8) presented the potential oncogenicity of *C. difficile* in a mouse model, to our knowledge, the prevalence of *C. difficile* detection in tumor and normal tissues from CRC resection has not previously been examined.

All *C. difficile*-positive tissues in our study displayed a low relative abundance of the organism, ranging from 0.005% to 0.3% of all assigned bacterial species, without a significant differential abundance or prevalence between tumor and paired normal tissues. Prior work using the same high-resolution taxonomic assignment method as our study (11) identified *C. difficile* at any abundance in the stool of ∼85% of patients clinically diagnosed with *C. difficile* infection (CDI, average abundance of 0.76-3.0%); in healthy control individuals examined in parallel, the percentage with any *C. difficile* detection dropped to ∼10% with a lower average relative abundance (0.008%). Our colon tissue detection rate in 38% of individuals and mean abundance of 0.04% in those individuals lies between these historic values characterizing active disease and asymptomatic carriage, suggesting a carriage burden potentially conducive to the persistent, low-level toxin production that murine studies have implicated in colonic tumorigenesis (8) or perhaps reinforcing the differential yields of mucosal versus stool samples for microbiome analysis as discussed below.

Notably, our limit of detection analysis demonstrates that even a cutoff depth of 9500 reads analyzed per sample results in only 58% sensitivity for the detection of any *C. difficile* read compared with our unrestricted analysis (mean of 19,501 reads per sample). While such limit of detection considerations may not be relevant in the analysis of other bacterial species suspected to be involved in colorectal tumorigenesis (such as pks+ *Escherichia coli*, which mouse model data suggest may be present at a relative abundance of 10-20% to influence tumorigenesis (16)), low level detection may be sufficient to promote tumorigenesis for keystone-type organisms such as *C. difficile*. Thus, our results suggest that limited read depth in 16S rRNA amplicon-based sequencing analyses of CRC samples risks missing potential contributors to CRC disease pathogenesis, a suggestion supported by recent evidence that low-level detection of asymptomatic, persistent *C. difficile* colonization in hospitalized patients is often persistent and associated with the later development of CDI (17).

We also noted a significantly higher rate of CRC biofilm detection in *C. difficile* positive individuals compared with those without detection of *C. difficile* in any tissues by 16S rRNA amplicon sequencing or culture. Bacterial biofilms are common on CRCs, can be oncogenic (4, 9, 18-20), and can be a contributor to *C. difficile* sporulation (affording antimicrobial and chemical endurance) (21), but no studies, to our knowledge, have yet drawn associations between CRC and *C. difficile* utilizing biofilm biology as a mediating factor. While the role of biofilm formation in the natural history of toxigenic *C. difficile* infections remains under-studied, recent work suggests that the synthesis of biofilms by this pathogen increases sporulation and decreases toxin production (22), consistent with our proposed model of persistent, low-level toxin production as a potential driver of colon oncogenesis.

The colon co-habitation of gram-negative rods (GNRs) appears to be an important influence on several key biologic processes related to the *C. difficile* lifecycle and may inform our hypothesis of colorectal tumorigenesis. We found enrichment of several GNRs belonging to the Proteobacteria phylum, including *Enterobacter, Klebsiella*, and *Parasutterella*, in the *C. difficile*-positive, biofilm-positive cohort compared with the *C. difficile*-negative, biofilm-positive group. Inflammatory GNRs, and particularly Enterobacteriaceae (such as *Klebsiella* and *Escherichia* species), have been shown through microbiome computational modeling studies to be enriched in individuals with recurrent *C. difficile* infection (rCDI) (23); putative mechanisms of how these GNRs may enable persistent/recurrent CDI included reductions in secondary bile acid synthesis and increases in aromatic amino acid metabolism. Our exploratory PICRUSt2 analysis provided limited signals suggesting metabolic differences between *C. difficile*-positive and -negative microbiota associated with CRC, findings requiring validation and development such as by metagenomic sequencing and/or direct metabolomic analyses.

Our study has several strengths, including being a large cohort of individuals with CRC and, most notably, our analysis of tissue rather than stool, including tumor and paired normal samples, by culture, PCR and/or nucleic acid sequencing. The combination of sequencing, culture, and molecular techniques to characterize *C. difficile* presence and linkage to CRC biofilms provides novel data. Prior data indicate that the human colon mucosa microbiota is either distinct from or only a subset of the fecal microbiota (24) and, thus, our analyses provide an initial human study aimed at beginning to translate our prior murine work (8). Prior data suggest that mucosal samples, rather than stool samples, generally contain useful information on the microbiota that is directly interactive with host tissue such as, in this study, the tumor microenvironment (25).

Our study also has limitations, the most crucial being the absence of a contemporaneous non-CRC control group with available colon tissue for parallel analysis, which limits any conclusions about the potential role of *C. difficile* in human CRC pathogenesis. Our inclusion of recent UMMC clinical microbiology laboratory data revealing low detection rates of *C. difficile* in fecal samples combined with limited prior literature on CDI in Malaysia suggest that detection of *C. difficile* in 38% of Malaysian individuals with CRC is unexpected. However, additional prospective studies with parallel controls and more detailed microbiota analyses (e.g., metagenomics, metabolomics, *C. difficile* strain analyses) are needed to validate our observations. Other notable limitations include lack of data on prior CDI history (including rCDI) in the enrollees and the marked heterogeneity in *C. difficile* detection between our sequencing cohorts (**Table S1**). While the latter is likely due to differences in DNA extraction and sequencing methods, it is important to note that over half of all samples analyzed were sequenced in the MAL3 cohort, which had the highest detection of *C. difficile* (57% of individuals with detection) and that this sequencing run occurred most recently and the increased detection may be consistent with improving sequencing and analysis technology. Importantly, although experimental data indicated that TcdB production is critical for *C. difficile*-induced oncogenesis (8), 16S rRNA amplicon sequencing cannot differentiate toxigenic from non-toxigenic *C. difficile*, requiring either alternate sequencing methodologies (e.g. metagenomic sequencing) or isolation of *C. difficile* strains by culture or molecular methods to identify toxigenic *C. difficile* in CRC tissues. We addressed this through targeted culture followed by molecular detection of *tcdB* and found that the *tcdB* gene was detected in 7 of the 10 *C. difficile* strains isolated, although we acknowledge the low culture yield. We suspect this is due to storage of samples at -80°C for >10 years.

In summary, low-abundance *C. difficile* was frequently detected in colon tissue in adults presenting for CRC resection in Malaysia and was associated with an increased likelihood of CRC-associated bacterial biofilms. These findings complement recent mouse studies implicating persistent colonic colonization with toxigenic *C. difficile* in colon tumorigenesis despite low relative abundance. These data support a potential unrecognized contribution of chronic, low-level toxigenic *C. difficile* colonization to mucus-invasive biofilm formation and CRC development in humans, a hypothesis requiring longitudinal studies of the natural history of *C. difficile* colonization that have not been conducted to date.

## SUPPLEMENTAL METHODS

### Selective Culture and Identification of *C. difficile* from Tissue Samples (Expanded)

3mm punch biopsies were collected from frozen tumor and normal tissue, mechanically homogenized, and resuspended in BHI (brain heart infusion) media supplemented with hemin (10 ug/mL) and vitamin K (5 ug/mL). The resulting slurries were plated on CCFA (Cycloserine-Cefoxitin Fructose Agar – Anaerobe Systems) and inoculated into CCMB-TAL (Cycloserine-Cefoxitin Mannitol Broth with Taurocholate Lysozyme Cysteine – Anaerobe Systems); cultures were incubated for 48-72 hours under anaerobic conditions (75% N_2_, 5% H_2_, 20% CO_2_) in an anaerobic chamber. Any color change in the CCMB-TAL tube triggered plating of the broth onto CCFA plates. From these two CCFA plates (1 directly plated, 1 subcultured first in CCMB-TAL), unique colonies were picked and spread onto new BHI plates for isolation (minimum 6 colonies/tumor) and incubated. After 48–72 hr, colonies were selected and swirled into sterile water, which was boiled to release bacterial DNA. This crude DNA preparation was screened by quantitative real-time PCR (qRT-PCR) with *C. difficile* species-specific 16S primers (forward, TTGAGCGATTTACTTCGGTAAAGA; reverse, CCATCCTGTACTGGCTCACCT) and probe (TAACGCGTGGGTAACCTACCCTGTA) (26), at a concentration of 0.2 μM, and with the following program: 50°C x 2 min; 95°C x 10 min; 40 cycles of 95°C x 30 sec, 58°C x 60 sec. Colonies were considered *C. difficile*-positive if the cycle threshold value was <30. Each *C. difficile*-positive colony was subcultured onto fresh BHI plates x 48 hr, and genomic DNA was extracted from pelleted bacteria using the ZR Quick-DNA Fecal/Soil Miniprep Kit (Zymo Research). Species identification of *C. difficile* was confirmed by Sanger sequencing of the full-length 16S rRNA gene (Azenta Life Sciences) for at least one colony from each individual who yielded a positive culture result.

### PCR for Toxin B Assay (Expanded)

*C. difficile* strains isolated by culture were analyzed for the presence of toxin genes (including *tcdA, tcdB, cdtA, cdtB*) utilizing a pan-toxin PCR as previously described (14). Briefly, DNA extracted from each isolated strain of *C. difficile* was amplified by PCR using primers for *tcdA, tcdB, cdtA*, and *cdtB*. PCR products were separated by electrophoresis on a 1.5% agarose gel, and the presence or absence of target bands at 410bp (*tcdB*) was assessed under UV illumination compared with the *C. difficile* reference strain M7404.

### Identification of Toxigenic *C. difficile* at UMMC (Control Population, Stool Samples)

Stool samples for detection of *C. difficile* toxins were stored at 2-8°C until C. DIFF QUIK CHEK COMPLETE^®^ was completed to detect *C. difficile* toxins A and B, generally within 24h. *C. difficile* stool cultures were performed by direct plating and alcohol shock culture methods. For direct plating, stool was inoculated on *C. difficile* agar and blood agar plates, with a metronidazole disc (5µg) placed on the first quadrant of both the plates, followed by anaerobic incubation at 35-37°C for 48 hours. For alcohol shock, an equal volume of 70% ethanol was added to the stool, vortex-mixed, held at room temperature for 1-hour followed by culture and incubation conditions as for direct plating. After 48 hours, suspicious colonies were identified using VITEK_®_ MS (bioMérieux SA, Marcy-I’Étoile, France).

## Data Availability

Raw sequences from MAL1, MAL2, and MAL3 were previously deposited in the NCBI SRA repository under BioProject accession nos. PRJNA325649, PRJNA325650, and PRJNA1195962, respectively. Any additional data or analyses are available by request.

## Data and code availability

Raw sequences from MAL1, MAL2, and MAL3 were previously deposited in the NCBI SRA repository under BioProject accession nos. PRJNA325649, PRJNA325650, and PRJNA1195962, respectively (9). Any additional data or analyses are available by request.

## FISH Analysis of Biofilms

Briefly, as previously described (9), sequential sections of tumor or normal tissues fixed in methcarn were stained with the Eub338 universal bacterial probe to enable detection of tissue biofilms. A biofilm was defined as a dense aggregation of mucus-invasive bacteria, with at least 20 bacteria within 1 μm of the epithelium, covering an expanse of at least 200 μm of the colon epithelial cell layer, in at least 1 of 3 screened areas.

## FIGURE/TABLE LEGENDS

**Supplemental Figure 1.**
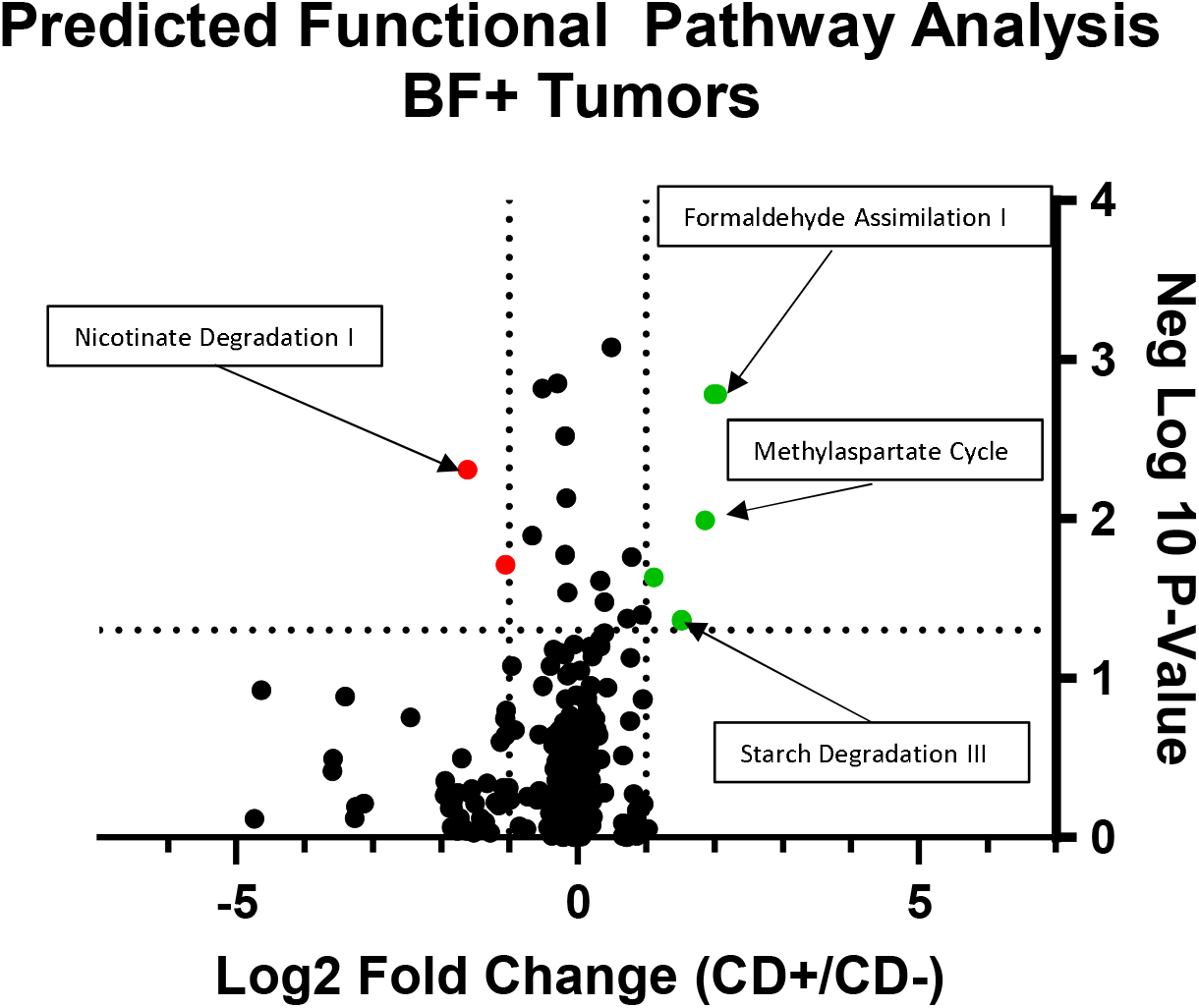
Community Metabolic Signatures of *C. difficile*-Positive Individuals. Volcano plot showing PICRUSt2 comparisons between the CD+ (*C. difficile* positive, biofilm positive) and CD-(*C. difficile* negative, biofilm positive) tumors as analyzed by Mann Whitney test, with p < 0.05 considered significant. Data is depicted as Log2-Fold Change (>0 indicates higher in CD+ cohort). Vertical dashed lines delineate -1 and +1, left to right. Horizontal dashed line indicates P value, 0.05 (displayed here as negative log10 of the p-value, with cutoff 1.33). Key pathways are labeled; see Supplemental Table 3 for full results of significant PICRUSt2 outputs.

**Supplemental Table 1.**
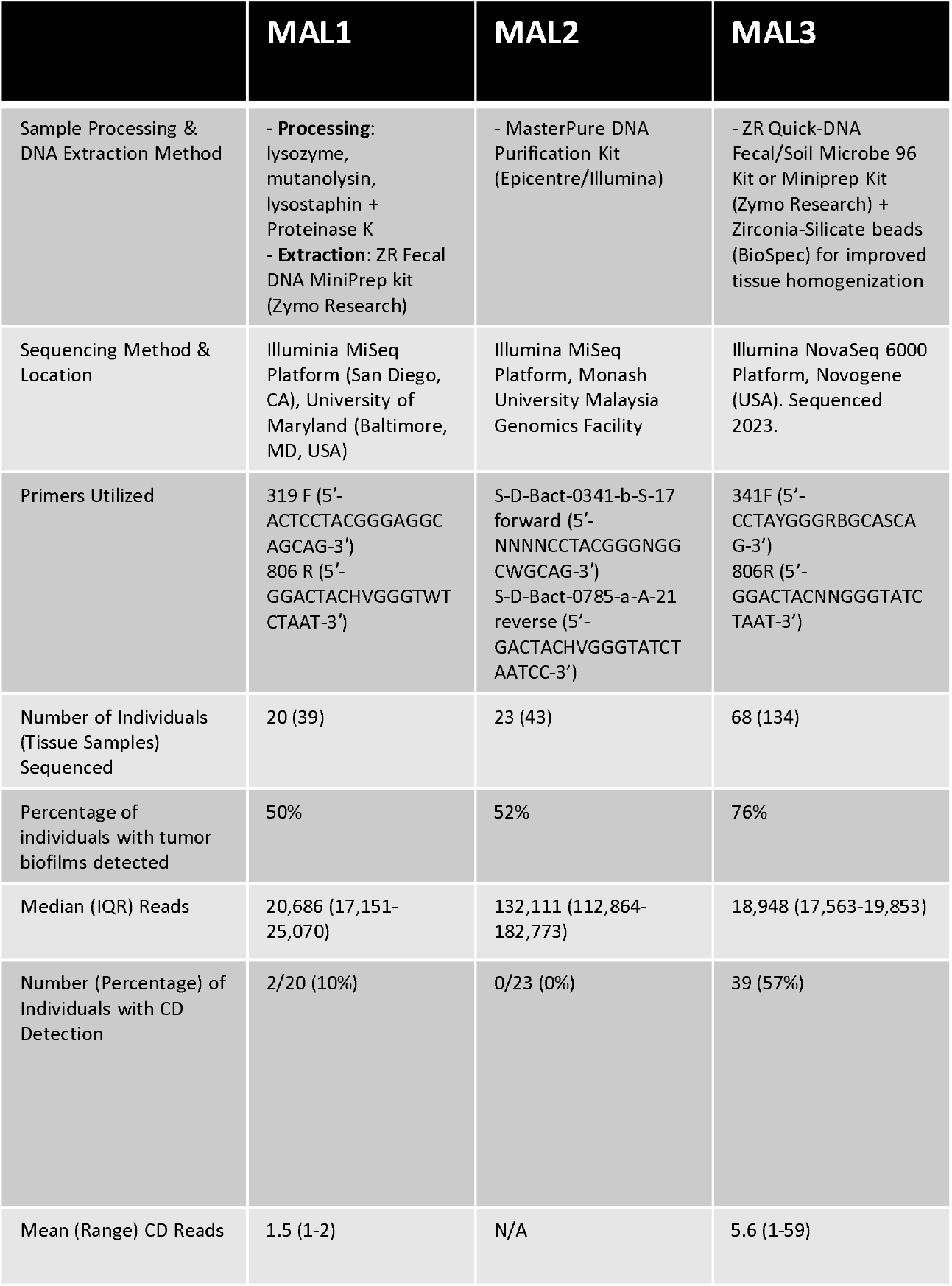
Sequencing Cohort Details and *C. difficile* Detection Rates. The three sub-cohorts of sequenced tissues (MAL1, MAL2, MAL3) differed in details of tissue processing; sequencing platform and location; primers utilized for amplification; the number of individuals and tissue samples analyzed; the frequency of bacterial biofilms detected; and detection rates of *C. difficile*. Note that some individuals had tumor tissue sequenced in one cohort and normal tissue sequenced in another, resulting in an apparent discordance in the number of individuals analyzed compared with those included in the study. The total number of tissues analyzed equals 216 (108 persons times two tissue samples sequenced per person).

**Supplemental Table 2.**
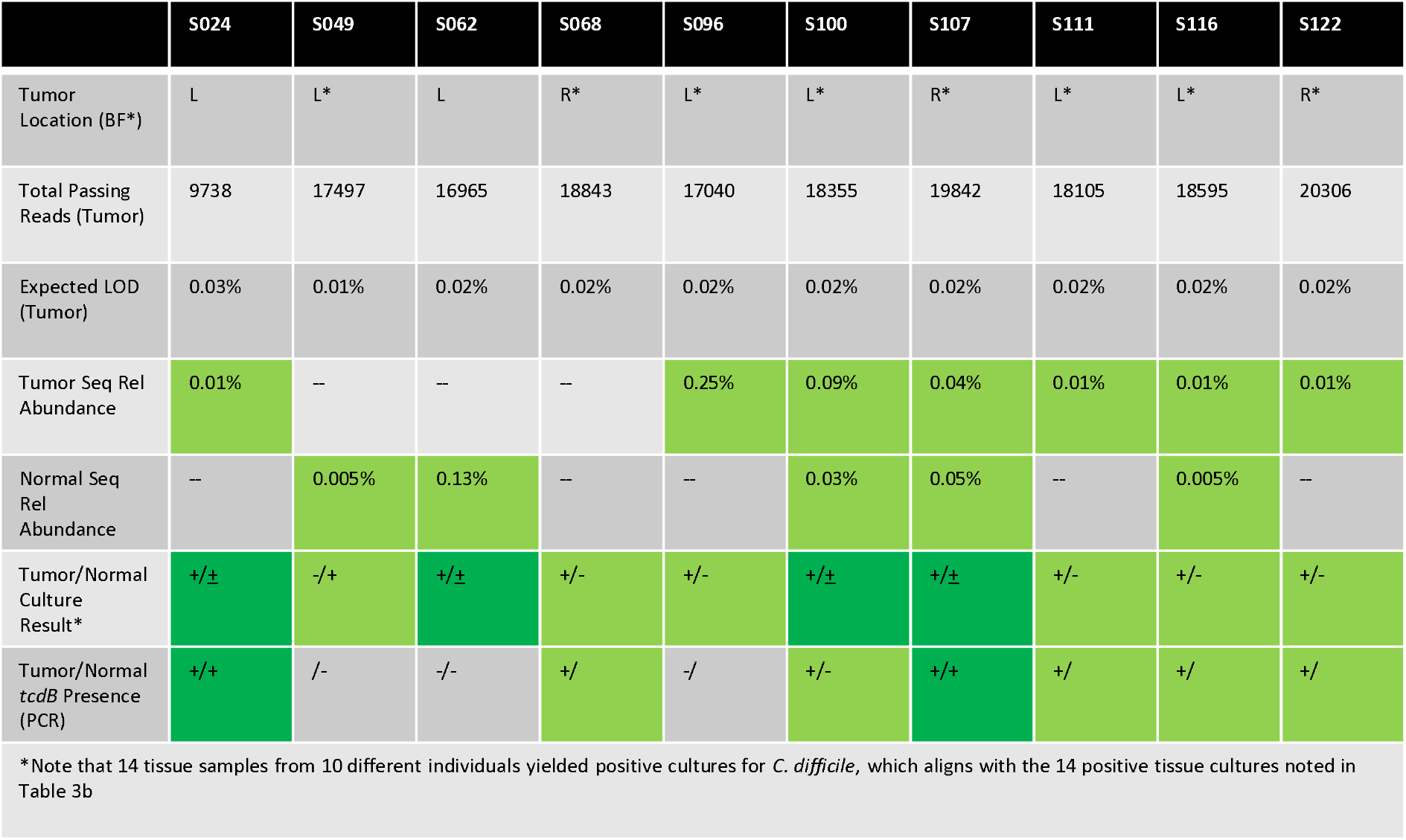
Tumor, Sequencing, Culture, and Molecular Characteristics of *C. difficile* Culture-Positive Individuals. Key tumor characteristics (left or right-sided location; biofilm present in individuals with asterisk); the total passing reads analyzed for the tumor tissue and expected limit of detection (LOD) for *C. difficile* based on calculations from Figure 1c; the relative abundance of *C. difficile* by sequencing in both tumor and normal tissue (Seq Rel Abundance); *C. difficile* culture success from both tissue types (“+” indicates positive culture for *C. difficile* in the respective tissue); and assessment of *tcdB* presence by PCR from each *C. difficile* isolate of the 10 individuals who were found to be culture-positive for *C. difficile* in at least one tissue type. Green shading represents any positivity; dark green shading depicts individuals with presumptive dual positive samples. Culture positive results with underlines were confirmed by qPCR but not Sanger Sequencing.

**Supplemental Table 3.** Significant PICRUSt2 Pathway Results. All significant pathways from PICRUSt2 analysis (see Supplemental Figure 1) are listed with a description and metabolic functional classification (derived from MetaCyc Ontology). Significance determined by pathways with a Log2-Fold change of greater than 1 (absolute value) and a p-value < 0.05 by Mann-Whitney U-Test [i.e., Q-value >1.33].

| Pathway | Description | Metabolic Function Group | Upregulated In | Q-Value | Log2 (Fold Change) |
| --- | --- | --- | --- | --- | --- |
| PWY_5529 (c1483) | superpathway of bacteriochlorophyll a biosynthesis | Secondary Metabolite Biosynthesis | CD+ | 2.78 | 2.04 |
| CHLOROPHYLL_SYN (c1324) | chlorophyllide a biosynthesis I (aerobic, light-dependent) | Secondary Metabolite Biosynthesis | CD+ | 2.78 | 1.99 |
| PWY_722 (c1630) | nicotinate degradation I | Aromatic Compound Degradation | CD- | 2.31 | -1.61 |
| PWY_1622 (c1433) | formaldehyde assimilation I (serine pathway) | Primary Metabolite Biosynthesis (Methanotrophic) | CD+ | 1.99 | 1.86 |
| DHGLUCONATE_PYR_CAT_PWY | glucose degradation (oxidative) | Carbohydrate Degradation | CD- | 1.71 | -1.05 |
| PWY_6728 (c1589) | methylaspartate cycle | Primary Metabolite Biosynthesis | CD+ | 1.64 | 1.12 |
| PWY_5531 (c1484) | chlorophyllide a biosynthesis II (anaerobic) | Secondary Metabolite Biosynthesis | CD+ | 1.36 | 1.52 |
| PWY_7159 (c1617) | chlorophyllide a biosynthesis III (aerobic, light independent) | Secondary Metabolite Biosynthesis | CD+ | 1.36 | 1.52 |
| PWY_6731 (c1590) | starch degradation III | Carbohydrate Degradation | CD+ | 1.34 | 1.64 |

**Supplemental Table 4.** *C. difficile* Fecal Culture-Positivity Results from non-CRC individuals evaluated in the UMMC clinical microbiology laboratory. All stool samples sent for toxigenic *C. difficile* testing at Universiti Malaya were assessed by culture and ToxinA/B EIAs (see Methods). Data from 2022 and 2023 are displayed with positive culture result and EIA status as described in Methods.

|  |  |  |
| --- | --- | --- |
| **2023**<br>N=1111 stools examined | **Number of strains** | **Positivity rate** |
| Culture positive for *C. difficile* | 51 | 4.59% |
| Toxin positive by EIA (A or B)* | 13 | 1.17% |
| **2022**<br>N=945 stools examined | **Number of strains** | **Positivity rate** |
| Culture positive for *C. difficile* | 68 | 7.20% |
| Toxin positive by EIA (A or B)* | 22 | 2.33% |

\*=Only culture-positive stools were analyzed for TcdA/B by EIA

## Author contributions

<u>Conceptualization</u> S.A. J.Q., C.L.S.; <u>Methodology</u>: S.A., J.Q., J.W., J.R.W, J.D., C.L.S.; <u>Software</u>: J.R.W.; <u>Formal Analysis</u>: S.A., J.Q., J.R.W.; <u>Validation</u>: S.A., J.Q., J.R.W.; <u>Investigation</u>: S.A., J.Q., Z.C., H.B., J.D., T.S., J.T.F., J.V., A.R., J.W., J.R.W., S.N.L.; <u>Resources</u>: S.A., J.Q., Z.C., J.V., J.D., C.L.S.; <u>Data Curation</u>: J.W., J.R.W., J.L.D., C.L.S.; Writing – Original Draft: S.A.; <u>Writing – Review & Editing</u>: all authors; <u>Visualization</u>: S.A., J.Q., J.R.W., J.D.; <u>Supervision</u>: J.Q., C.L.S.; <u>Project Administration</u>: J.W., C.L.S.; <u>Funding Acquisition</u>: S.A., J.Q., C.L.S.

## References

1. Siegel RL, Miller KD, Fedewa SA, Ahnen DJ, Meester RGS, Barzi A, et al. Colorectal cancer statistics, 2017. CA Cancer J Clin. 2017;67(3):177–93. Epub 20170301. doi: 10.3322/caac.21395. PubMed PMID: 28248415.

2. Siegel RL, Wagle NS, Jemal A. Leading Cancer Deaths in People Younger Than 50 Years. JAMA. 2026. Epub 20260122. doi: 10.1001/jama.2025.25467. PubMed PMID: 41569583; PubMed Central PMCID: PMC12828652.

3. White MT, Sears CL. The microbial landscape of colorectal cancer. Nat Rev Microbiol. 2024;22(4):240–54. Epub 20231004. doi: 10.1038/s41579-023-00973-4. PubMed PMID: 37794172.

4. Drewes JL, White JR, Dejea CM, Fathi P, Iyadorai T, Vadivelu J, et al. High-resolution bacterial 16S rRNA gene profile meta-analysis and biofilm status reveal common colorectal cancer consortia. NPJ Biofilms Microbiomes. 2017;3:34. Epub 20171129. doi: 10.1038/s41522-017-0040-3. PubMed PMID: 29214046; PubMed Central PMCID: PMC5707393.

5. Pleguezuelos-Manzano C, Puschhof J, Rosendahl Huber A, van Hoeck A, Wood HM, Nomburg J, et al. Mutational signature in colorectal cancer caused by genotoxic pks. Nature. 2020;580(7802):269–73. Epub 20200227. doi: 10.1038/s41586-020-2080-8. PubMed PMID: 32106218; PubMed Central PMCID: PMC8142898.

6. Kostic AD, Gevers D, Pedamallu CS, Michaud M, Duke F, Earl AM, et al. Genomic analysis identifies association of Fusobacterium with colorectal carcinoma. Genome Res. 2012;22(2):292–8. Epub 20111018. doi: 10.1101/gr.126573.111. PubMed PMID: 22009990; PubMed Central PMCID: PMC3266036.

7. Boleij A, Hechenbleikner EM, Goodwin AC, Badani R, Stein EM, Lazarev MG, et al. The Bacteroides fragilis toxin gene is prevalent in the colon mucosa of colorectal cancer patients. Clin Infect Dis. 2015;60(2):208–15. Epub 20141009. doi: 10.1093/cid/ciu787. PubMed PMID: 25305284; PubMed Central PMCID: PMC4351371.

8. Drewes JL, Chen J, Markham NO, Knippel RJ, Domingue JC, Tam AJ, et al. Human Colon Cancer– Derived Clostridioides difficile Strains Drive Colonic Tumorigenesis in Mice. Cancer Discovery. 2022;12(8):1873–85. doi: 10.1158/2159-8290.CD-21-1273.

9. Queen J, Cing Z, Minsky H, Nandi A, Southward T, Ferri J, et al. Fusobacterium nucleatum is enriched in invasive biofilms in colorectal cancer. NPJ Biofilms Microbiomes. 2025;11(1):81. Epub 20250520. doi: 10.1038/s41522-025-00717-7. PubMed PMID: 40394001; PubMed Central PMCID: PMC12092649.

10. Grim CJ, Daquigan N, Lusk Pfefer TS, Ottesen AR, White JR, Jarvis KG. High-Resolution Microbiome Profiling for Detection and Tracking of. Front Microbiol. 2017;8:1587. Epub 20170818. doi: 10.3389/fmicb.2017.01587. PubMed PMID: 28868052; PubMed Central PMCID: PMC5563311.

11. Daquigan N, Seekatz AM, Greathouse KL, Young VB, White JR. High-resolution profiling of the gut microbiome reveals the extent of. NPJ Biofilms Microbiomes. 2017;3:35. Epub 20171205. doi: 10.1038/s41522-017-0043-0. PubMed PMID: 29214047; PubMed Central PMCID: PMC5717231.

12. Langille MG, Zaneveld J, Caporaso JG, McDonald D, Knights D, Reyes JA, et al. Predictive functional profiling of microbial communities using 16S rRNA marker gene sequences. Nat Biotechnol. 2013;31(9):814–21. Epub 20130825. doi: 10.1038/nbt.2676. PubMed PMID: 23975157; PubMed Central PMCID: PMC3819121.

13. Sheikh IA, Bianchi-Smak J, Laubitz D, Schiro G, Midura-Kiela MT, Besselsen DG, et al. Transplant of microbiota from Crohn’s disease patients to germ-free mice results in colitis. Gut Microbes. 2024;16(1):2333483. Epub 20240327. doi: 10.1080/19490976.2024.2333483. PubMed PMID: 38532703; PubMed Central PMCID: PMC10978031.

14. Persson S, Torpdahl M, Olsen KE. New multiplex PCR method for the detection of Clostridium difficile toxin A (tcdA) and toxin B (tcdB) and the binary toxin (cdtA/cdtB) genes applied to a Danish strain collection. Clin Microbiol Infect. 2008;14(11):1057–64. doi: 10.1111/j.1469-0691.2008.02092.x. PubMed PMID: 19040478.

15. Collins DA, Hawkey PM, Riley TV. Epidemiology of Clostridium difficile infection in Asia. Antimicrob Resist Infect Control. 2013;2(1):21. Epub 20130701. doi: 10.1186/2047-2994-2-21. PubMed PMID: 23816346; PubMed Central PMCID: PMC3718645.

16. Arthur JC, Perez-Chanona E, Mühlbauer M, Tomkovich S, Uronis JM, Fan TJ, et al. Intestinal inflammation targets cancer-inducing activity of the microbiota. Science. 2012;338(6103):120–3. Epub 20120816. doi: 10.1126/science.1224820. PubMed PMID: 22903521; PubMed Central PMCID: PMC3645302.

17. Miles-Jay A, Snitkin ES, Lin MY, Shimasaki T, Schoeny M, Fukuda C, et al. Longitudinal genomic surveillance of carriage and transmission of Clostridioides difficile in an intensive care unit. Nat Med. 2023;29(10):2526–34. Epub 20230918. doi: 10.1038/s41591-023-02549-4. PubMed PMID: 37723252; PubMed Central PMCID: PMC10579090.

18. Dejea CM, Wick EC, Hechenbleikner EM, White JR, Mark Welch JL, Rossetti BJ, et al. Microbiota organization is a distinct feature of proximal colorectal cancers. Proc Natl Acad Sci U S A. 2014;111(51):18321–6. Epub 20141208. doi: 10.1073/pnas.1406199111. PubMed PMID: 25489084; PubMed Central PMCID: PMC4280621.

19. Dejea CM, Fathi P, Craig JM, Boleij A, Taddese R, Geis AL, et al. Patients with familial adenomatous polyposis harbor colonic biofilms containing tumorigenic bacteria. Science. 2018;359(6375):592–7. Epub 20180201. doi: 10.1126/science.aah3648. PubMed PMID: 29420293; PubMed Central PMCID: PMC5881113.

20. Tomkovich S, Dejea CM, Winglee K, Drewes JL, Chung L, Housseau F, et al. Human colon mucosal biofilms from healthy or colon cancer hosts are carcinogenic. J Clin Invest. 2019;129(4):1699–712. Epub 20190311. doi: 10.1172/JCI124196. PubMed PMID: 30855275; PubMed Central PMCID: PMC6436866.

21. Dawson LF, Peltier J, Hall CL, Harrison MA, Derakhshan M, Shaw HA, et al. Extracellular DNA, cell surface proteins and c-di-GMP promote biofilm formation in Clostridioides difficile. Sci Rep. 2021;11(1):3244. Epub 20210205. doi: 10.1038/s41598-020-78437-5. PubMed PMID: 33547340; PubMed Central PMCID: PMC7865049.

22. Purcell EB, McKee RW, Courson DS, Garrett EM, McBride SM, Cheney RE, et al. A Nutrient-Regulated Cyclic Diguanylate Phosphodiesterase Controls Clostridium difficile Biofilm and Toxin Production during Stationary Phase. Infect Immun. 2017;85(9). Epub 20170818. doi: 10.1128/IAI.00347-17. PubMed PMID: 28652311; PubMed Central PMCID: PMC5563577.

23. Henson MA. Computational modeling of the gut microbiota reveals putative metabolic mechanisms of recurrent Clostridioides difficile infection. PLoS Comput Biol. 2021;17(2):e1008782. Epub 20210222. doi: 10.1371/journal.pcbi.1008782. PubMed PMID: 33617526; PubMed Central PMCID: PMC7932513.

24. Shalon D, Culver RN, Grembi JA, Folz J, Treit PV, Shi H, et al. Profiling the human intestinal environment under physiological conditions. Nature. 2023;617(7961):581–91. Epub 20230510. doi: 10.1038/s41586-023-05989-7. PubMed PMID: 37165188; PubMed Central PMCID: PMC10191855.

25. Valciukiene J, Strupas K, Poskus T. Tissue vs. Fecal-Derived Bacterial Dysbiosis in Precancerous Colorectal Lesions: A Systematic Review. Cancers (Basel). 2023;15(5). Epub 20230304. doi: 10.3390/cancers15051602. PubMed PMID: 36900392; PubMed Central PMCID: PMC10000868.

26. Rinttilä T, Kassinen A, Malinen E, Krogius L, Palva A. Development of an extensive set of 16S rDNA-targeted primers for quantification of pathogenic and indigenous bacteria in faecal samples by real-time PCR. J Appl Microbiol. 2004;97(6):1166–77. doi: 10.1111/j.1365-2672.2004.02409.x. PubMed PMID: 15546407.

